# Regression Tree Modelling to Predict Total Average Extra Costs in Household Spending During COVID-19 Pandemic

**DOI:** 10.1101/2021.01.15.21249732

**Authors:** Nesma Lotfy

## Abstract

**Background:** Prevention of coronavirus (COVID-19) regarding households has many aspects, such as buying masks, hand sanitizer, face shield, and many others. As a result of buying the previous items, the household spending per month will be increase during the COVID-19 pandemic period.

**Aims:** To calculate the average costs of each extra item involved in households spending during COVID-19 pandemic and to predict the total average extra costs spending by households.

**Methods:** A cross-sectional study was conducted at High Institute of Public Health (HIPH), University Alexandria. Exponential snowball sampling was used to recruit students at HIPH and their friends. Trimming costs was done to remove extreme low and high values. A regression tree modelling was implemented to predict the total extra costs spending during COVID-19 pandemic.

**Results:** Most of the respondents were female (81%) and aged between 30 and 40 (56.3%). About 63.1% of families had the same monthly income while 35.4% had a decrease in monthly income. A significant reduction in days of leaving home before and after COVID-19 pandemic was observed (before; mean= 5.86, after; mean = 4.66, *P*=0.000). The extra spending in grocery was the dominated item compared to other items (mean = 707.2, SD = 530.7). Regarding regression tree, the maximum average extra costs due to COVID-19 pandemic was 1386 L.E/month (around 88.23$/month) while the minimum average extra costs was 217 L.E/month (around 13.81$/month).

**Conclusions:** The effect of COVID-19 pandemic in households spending varies largely between households, it depends on what they do to prevent COVID-19.

**\*Hint: Convert form L.E to dollar performed according to price of dollar at 18-12-2020**

## Introduction

The coronavirus disease-2019 (COVID-19) is an ongoing worldwide pandemic caused by severe acute respiratory syndrome coronavirus 2 (SARS CoV-2). The virus was started in Wuhan, China but then it spread to all over the world. On January 2020, the World Health Organization declared that the COVID-19 outbreak as a global health emergency. Later, it declared it as a global pandemic (1).

The COVID-19 virus infects all ages. However, older aged people and patients with underlying medical conditions (such as hypertension, diabetes, cardiovascular disease, chronic respiratory disease, chronic kidney disease, immunocompromised status, cancer, pregnancy, smoking and obesity) are at a higher risk of severe COVID-19 disease. Severe COVID-19 disease causes hospitalization, admission to the ICU, intubation or mechanical ventilation, or death (2).

Prevention of coronavirus (COVID-19) is through regular hand washing and covering mouth and nose during sneezing or coughing. Avoidance of closed spaces and crowded places is also crucial for prevention of viral spread (3).

From our daily life in Alexandria, Egypt, there are also many undertaken promotions to prevent COVID-19 such as taking vitamins (vitamin C, zinc, …), conduct periodic examination, use taxi or uber instead of public transportation, increase the school expenses due to COVID-19 and many others. Thus, the main objective of the current study is to calculate the average extra costs of each item involved in households spending and to build a model to predict the total average extra costs spending by households due to COVID-19 pandemic.

## Methodology

### Study design

A cross-sectional study was conducted from October 2020 to November 2020 at High Institute of Public Health (HIPH), University Alexandria.

### Sampling design

Exponential snowball sampling technique was done to recruit HIPH students, their friends and family.

### Data collection methods and tools

A pre-design structure questionnaire was created to measure the impact of COVID-19 situation on the economics of Egyptian households. Content validity was assessed by three professors at HIPH, they agreed about the main items that must be asked to measure the average extra costs due to COVID-19 pandemic which are masks, face shield, treatment of covid-19, hand wash, chlorine, hand sanitizer, vitamins, periodic examination, transportation, delivery service, grocery spending, school expenses, and university expenses.

### Data management

The questionnaire was completed by a total of 280 respondents, about 17 (6.1%) individuals submitted more than one application so the duplicates were excluded from the study, and the remaining were 263 respondents. The data was collected online using google form.

### ✥ Data cleaning

After collection data, it is coded and fed to SPSS (statistical package for the social sciences (IBM® SPSS® Statistics version 25.0).

The costs were trimmed to remove the extreme low and extreme high values, so only values between 10% percentile and 90% percentile were entered in calculation.

### ✥ Calculation of average extra costs for each item per month

- Average cost of masks per month if they bought.
- Average cost of face shield for children if they bought.
  - It was assumed that the face shield will bought every 3 months), So the average cost of face shield per month was calculated as follows:
    - (Cost of face shield ∗ number of face shield bought)/3
- Average cost of treatment for COVID-19 if someone infected in the family
  - It was assumed that the individual that was infected will be infected only one time per year, so the average cost of treatment per month was calculated as follows:
    - (Cost of treatment ∗ number of individual infected)/12
- Average extra cost of hand wash per month.
- Average extra cost of chlorine per month.
- Average cost of hand sanitizer if they bought
  - It was calculated as follows:
    - Cost of hand sanitizer ∗ number of bottles used per month.
- Average costs of taking vitamins per month if they bought
- Average costs of mouth gargle if they bought
  - It was assumed that 2 bottles will be used per month, so average costs of mouth gargle per month was calculated as follows:
    - Cost of one bottle ∗ 2
- Average cost of periodic examination per month.
- Average extra cost of using Uber or Taxi instead of public transportation if they use.
  - Increasing cost calculated as: the cost of using uber or taxi per days for one person in the family subtracting by the cost of public transportation (around 15 L.E per day)
  - The increasing cost was multiplied by 15 days (average days of coming out from house)
- Average extra cost of using delivery service if they use
  - If they were usually use delivery service before COVID-19 situation and then stops, it was calculated as follows:
    - (delivery service fees (max 20 L.E) ∗ 15 days
      - If they were usually not use delivery service before COVID-19 situation and then use it
    - (delivery service fees (max 20 L.E) ∗ 15 days
- Average extra cost of grocery spending per month
- Average extra costs of school expenses for children per month
  - It was calculated as follows:
    - Extra costs for all children /12
  - Average extra costs of university expenses for students per month
    - It was calculated as follows:
      - (Extra costs for one student ∗ number of students) / 12

### ✥ Calculation of Average extra total cost

- Total cost was calculated as follows:
  - First: calculate the average extra cost for each item according to the whole population, this was done by multiplying each item cost for each individual by the proportion of the population who paid extra cost for it. For example: if individual_1 paid for masks 100 L.E, and 95% of population uses masks, then the cost for this individual will be 100∗.95= 95 L.E
  - Second: sum all average extra costs for all items, to get the total extra costs for each individual.

### Statistical analysis

#### ➢ Descriptive analysis

Descriptive statistics were calculated using min, max, mean, and standard deviation (SD) for the numeric data and frequency, and percent for categorical data.

Mann-Whitney test was conducted to test the significant difference between household leaving days before and after COVID-19 pandemic.

#### ➢ Regression tree modelling

Regression tree modelling technique using CART (Classification and Regression Trees) algorithm (4) was implemented to build a prediction model in terms of if-else rules to predict the total average extra costs spend per month during COVID-19 period based on its items. The regression tree has root nodes and leaf nodes. The root nodes represent a single input value (x) and a split point on that variable, while the leaf nodes represent the output variable (y). The CART algorithm involving selecting the input variables is based on a greedy algorithm which depends on minimization of sum square errors. The analysis was carried out using the R Statistical Environment (5) through rpart package (6).

## Results

Concerning to characteristics of respondents, more than half ages between 30 and 40 (56.3%), females (81%), married (62.4%) and from Alexandria (71.5%) (Table 1).

**Table 1:** Demographic characteristics of the respondents.

| Variable | N (%) |
| --- | --- |
|  | **N=263** |
| **Age** |  |
| 21- | 67(25.5) |
| 30- | 148(56.3) |
| 40+ | 48(18.3) |
| **Gender** |  |
| Male | 50(19.0) |
| Female | 213(81.0) |
| **Marital status** |  |
| Married | 164(62.4) |
| Single and live with parents | 74(28.1) |
| Single (live alone) | 8(3.0) |
| Widow or divorce | 17(6.5) |
| **Number of individuals in a family** |  |
| 1 | 13(4.9) |
| 2 | 21(8.0) |
| 3 | 57(21.7) |
| 4 | 90(34.2) |
| 5 | 70(26.6) |
| 6+ | 12(4.5) |
| **Residence** |  |
| Alexandria | 188(71.5) |
| Bahira | 26(9.9) |
| Other | 49(18.6) |

Regarding to COVID-19 impact on households spending, around 63.1% of families had the same monthly income, 1.5% had an increase with a mean of 475(234.2), and 35.4% had a decrease with a mean of 1788.31(962.9). About 87.5% of families had no persons who lost their jobs due to COVID-19, 11% had only one person while 1.5% had two persons. Concerning to change of household spending, 35.4% had no change, 49.8% had increase in household spending by a mean (SD) of 1215.65(560.83), and 14.8% had a decrease in spending by a mean (SD) of 1702.7(989.24) (Table 2).

**Table 2:** COVID-19 impact on households spending.

| Variable | N (%) | Costs (L.E)/Month |  |  |  |
| --- | --- | --- | --- | --- | --- |
|  | N=263 | N | Min | Max | Mean (SD) |
| Is the monthly income affected by the Corona pandemic? |  |  |  |  |  |
| No change | 166(63.1) |  |  |  |  |
| Increase | 4(1.5) | 4 | 300 | 800 | 475(234.2) |
| Decrease | 93(35.4) | 77 | 500 | 4000 | 1788.31(962.9) |
| How many members of your family lost their job due to the Corona pandemic |  |  |  |  |  |
| 0 | 230(87.5) |  |  |  |  |
| 1 | 29(11.0) |  |  |  |  |
| 2 | 4(1.5) |  |  |  |  |
| How much did you spend during one month on family expenses such as food, medicine and utilities (before the outbreak of the epidemic in Egypt, March 2020)? |  | 230 | 2000 | 9000 | 3769.57(1583.6) |
| Has there been a change in household spending during the outbreak in Egypt? |  |  |  |  |  |
| No change | 93(35.4) |  |  |  |  |
| Increase | 131(49.8) | 115 | 500 | 2000 | 1215.65(560.83) |
| Decrease | 39(14.8) | 37 | 500 | 4000 | 1702.7(989.24) |

Concerning to COVID-19 impact on households staying, there is a significant observed reduction in days of leaving home before and after COVID-pandemic for both the respondents (before; mean= 5.86, after; mean = 4.66, *P*=0.000) and for his family members (before; mean= 5.15, after; mean = 3.89, P=0.000) (Table 3).

**Table 3:** COVID-19 impact on households staying.

| Variables | Before covid-19 | After covid-19 | Sig |
| --- | --- | --- | --- |
| How many days do you leave the house during the week | 5.86(1.3) | 4.66(1.9) | 0.000 |
| For family members, the average days that they leave the house during the week | 5.15(1.7) | 3.89(2.04) | 0.000 |

Table 4 represents the average extra costs for each item during COVID-19 pandemic. Most of extra spending was for grocery (mean = 707.2, SD = 530.7), the second one was for using taxies or Ubers instead of public transportation (mean = 488.9, SD = 401.6), the third one was the cost of treatment for COVID-19 patients (mean = 348.4, SD = 344.5), the fourth one was for periodic examination (mean = 338.3, SD= 160.9). Figure 1 shows the mean extra costs of all items with SD as error bars.

**Table 4:**
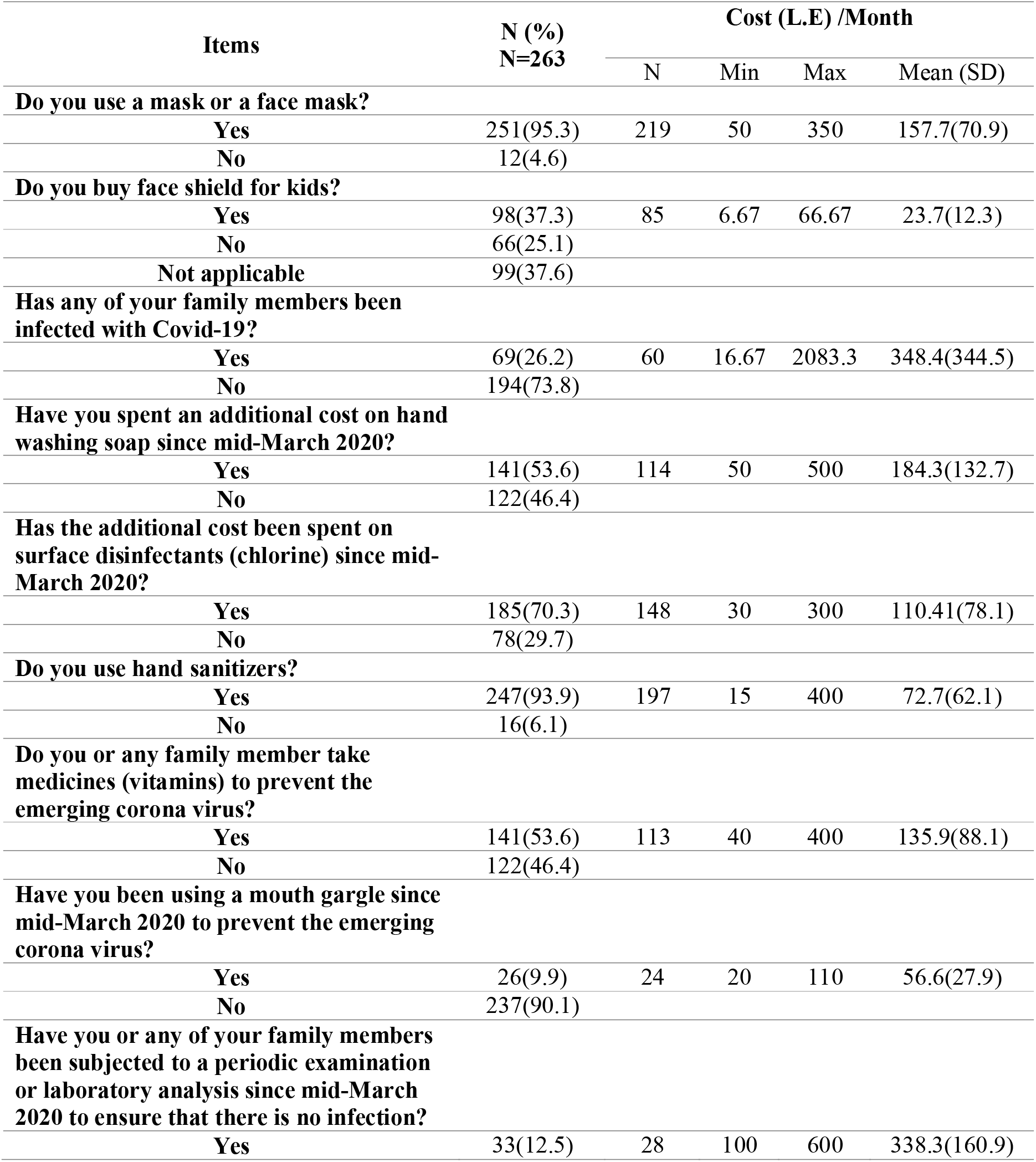
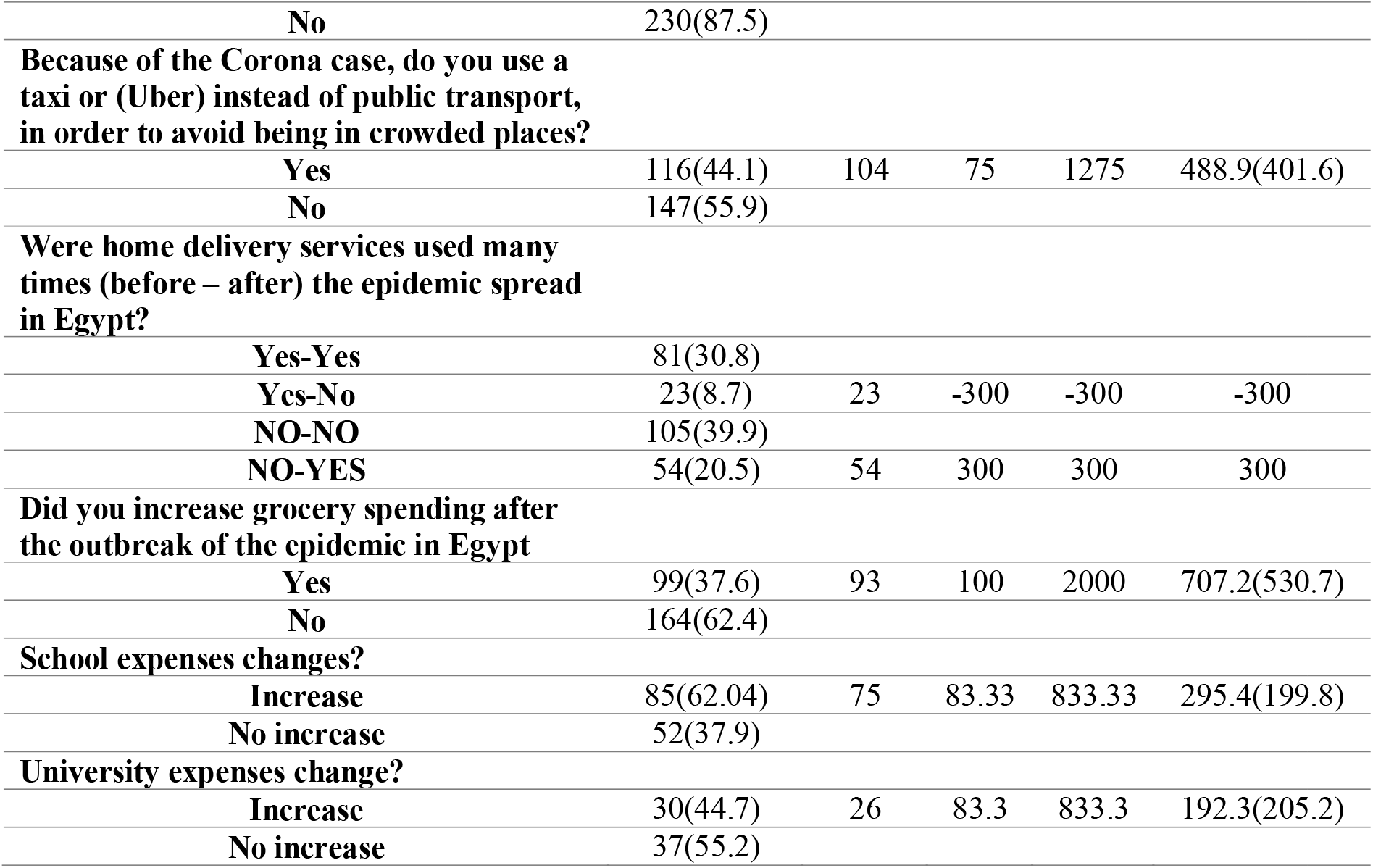
Average extra costs of each item during COVID-19 pandemic.

| Items | N (%)<br>N=263 | Cost (L.E) /Month |  |  |  |
| --- | --- | --- | --- | --- | --- |
|  |  | N | Min | Max | Mean (SD) |
| **Do you use a mask or a face mask?** |  |  |  |  |  |
| Yes | 251(95.3) | 219 | 50 | 350 | 157.7(70.9) |
| No | 12(4.6) |  |  |  |  |
| **Do you buy face shield for kids?** |  |  |  |  |  |
| Yes | 98(37.3) | 85 | 6.67 | 66.67 | 23.7(12.3) |
| No | 66(25.1) |  |  |  |  |
| Not applicable | 99(37.6) |  |  |  |  |
| **Has any of your family members been infected with Covid-19?** |  |  |  |  |  |
| Yes | 69(26.2) | 60 | 16.67 | 2083.3 | 348.4(344.5) |
| No | 194(73.8) |  |  |  |  |
| **Have you spent an additional cost on hand washing soap since mid-March 2020?** |  |  |  |  |  |
| Yes | 141(53.6) | 114 | 50 | 500 | 184.3(132.7) |
| No | 122(46.4) |  |  |  |  |
| **Has the additional cost been spent on surface disinfectants (chlorine) since mid-March 2020?** |  |  |  |  |  |
| Yes | 185(70.3) | 148 | 30 | 300 | 110.41(78.1) |
| No | 78(29.7) |  |  |  |  |
| **Do you use hand sanitizers?** |  |  |  |  |  |
| Yes | 247(93.9) | 197 | 15 | 400 | 72.7(62.1) |
| No | 16(6.1) |  |  |  |  |
| **Do you or any family member take medicines (vitamins) to prevent the emerging corona virus?** |  |  |  |  |  |
| Yes | 141(53.6) | 113 | 40 | 400 | 135.9(88.1) |
| No | 122(46.4) |  |  |  |  |
| **Have you been using a mouth gargle since mid-March 2020 to prevent the emerging corona virus?** |  |  |  |  |  |
| Yes | 26(9.9) | 24 | 20 | 110 | 56.6(27.9) |
| No | 237(90.1) |  |  |  |  |
| **Have you or any of your family members been subjected to a periodic examination or laboratory analysis since mid-March 2020 to ensure that there is no infection?** |  |  |  |  |  |
| Yes | 33(12.5) | 28 | 100 | 600 | 338.3(160.9) |
| No | 230(87.5) |  |  |  |  |
| **Because of the Corona case, do you use a taxi or (Uber) instead of public transport, in order to avoid being in crowded places?** |  |  |  |  |  |
| Yes | 116(44.1) | 104 | 75 | 1275 | 488.9(401.6) |
| No | 147(55.9) |  |  |  |  |
| **Were home delivery services used many times (before – after) the epidemic spread in Egypt?** |  |  |  |  |  |
| Yes-Yes | 81(30.8) |  |  |  |  |
| Yes-No | 23(8.7) | 23 | -300 | -300 | -300 |
| NO-NO | 105(39.9) |  |  |  |  |
| NO-YES | 54(20.5) | 54 | 300 | 300 | 300 |
| **Did you increase grocery spending after the outbreak of the epidemic in Egypt** |  |  |  |  |  |
| Yes | 99(37.6) | 93 | 100 | 2000 | 707.2(530.7) |
| No | 164(62.4) |  |  |  |  |
| **School expenses changes?** |  |  |  |  |  |
| Increase | 85(62.04) | 75 | 83.33 | 833.33 | 295.4(199.8) |
| No increase | 52(37.9) |  |  |  |  |
| **University expenses change?** |  |  |  |  |  |
| Increase | 30(44.7) | 26 | 83.3 | 833.3 | 192.3(205.2) |
| No increase | 37(55.2) |  |  |  |  |

**Figure 1:**
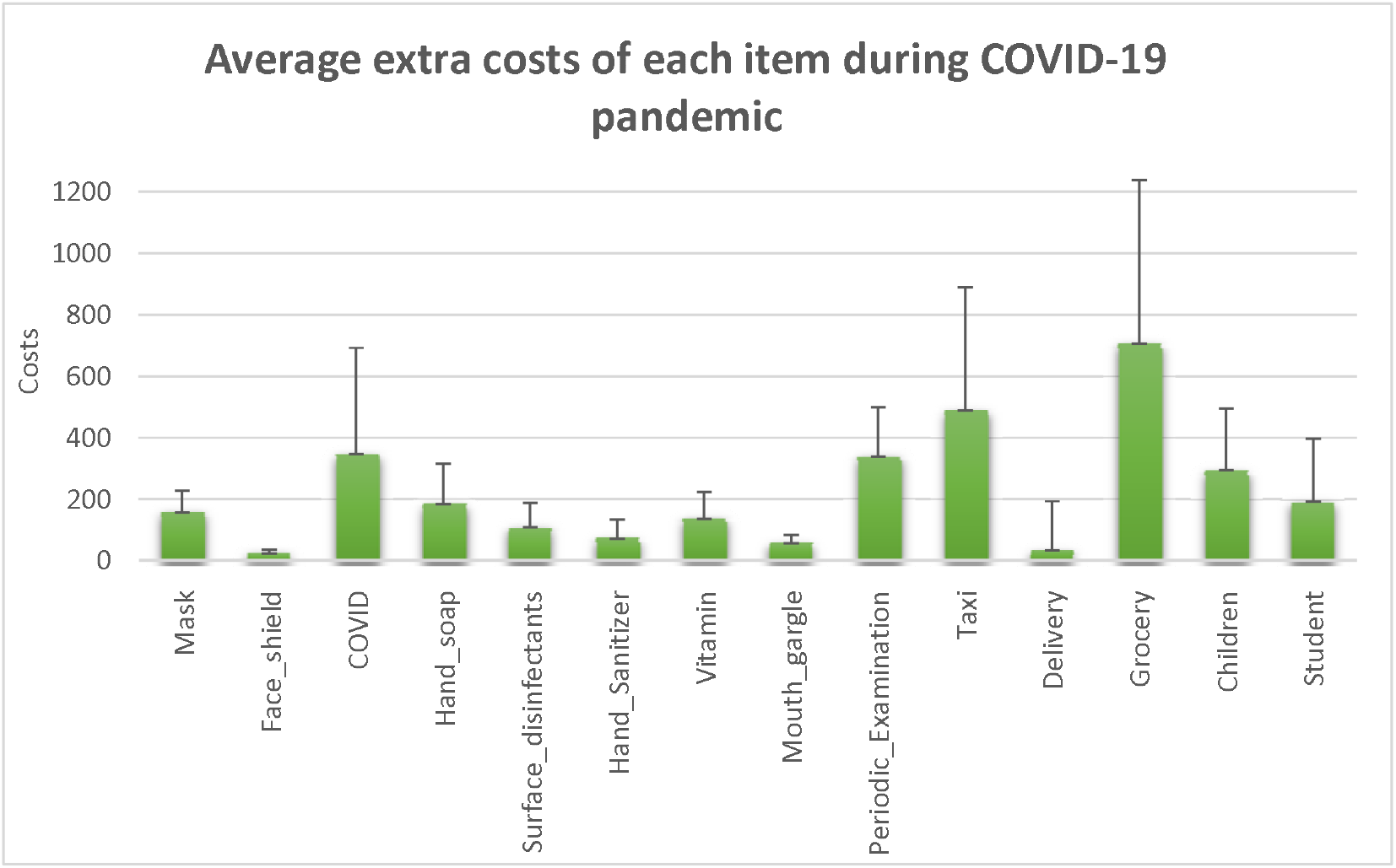
Average extra costs of each item during COVID-19 pandemic.

The regression tree selected only six items from the 14 items to build the tree. These items are Grocery, Surface disinfectants, Taxi, Vitamin, Face shield, and Children. The tree has 8 leaves with a main root which is Grocery items (Figure 2).

**Figure 2:**
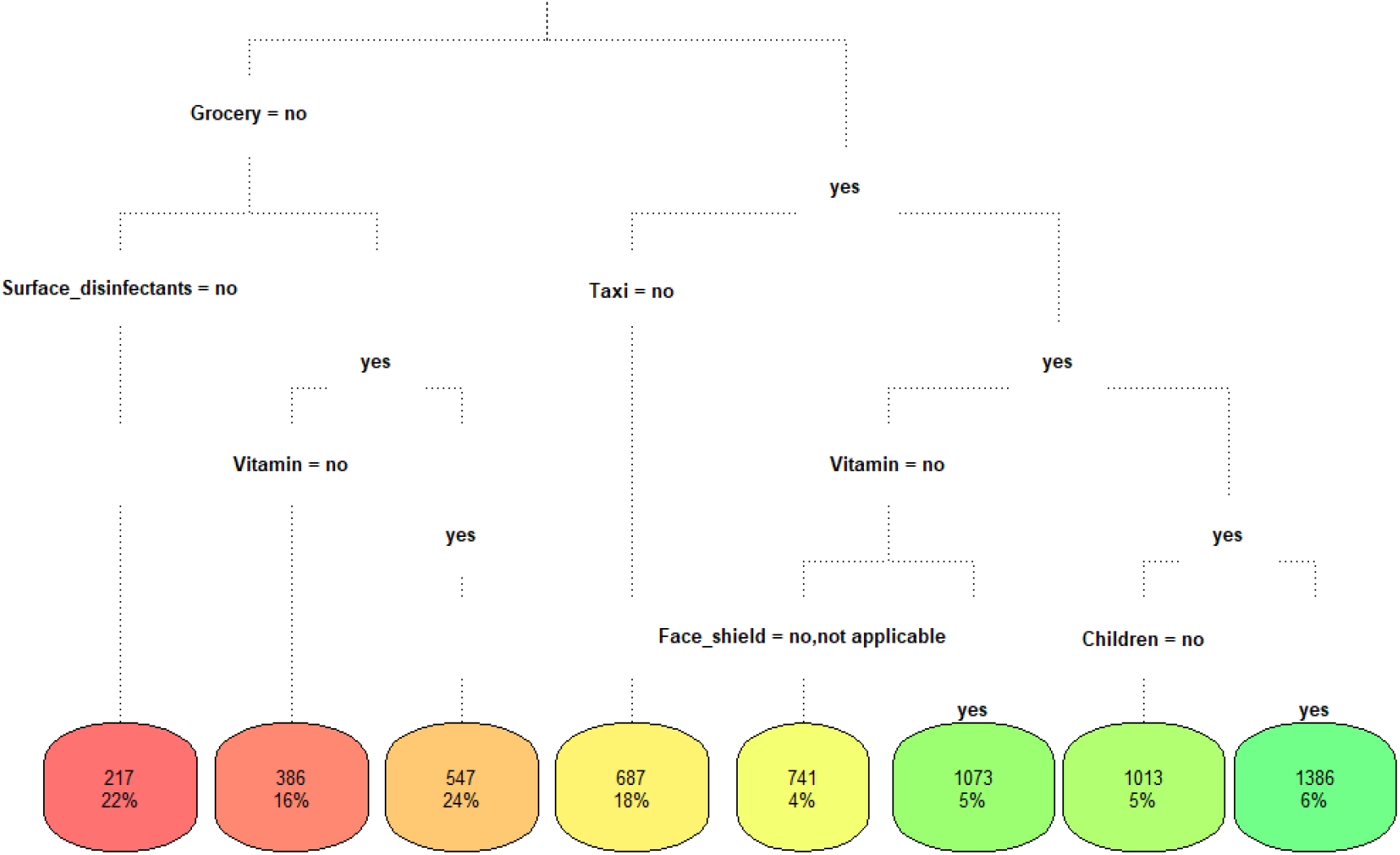
Regression tree modelling for predicting the average extra costs per month.

Table 5 shows the decision rules of the regression tree, it can be read as follows, for example: if you have no increase in Grocery spending and have no increase in surface disinfectants spending then the average extra cost of spending per month is 217 L.E. Thus, the minimum average extra cost spending per month is 217 L.E. and the maximum average extra cost spending per month is 1386 L.E.

**Table 5:** Decision rules of the regression tree.

| **Rules** | **Cost**<br>**(L.E)/Month** |
| --- | --- |
| When Grocery is no & Surface disinfectants is no | **217** |
| When Grocery is no & Vitamin is no & Surface disinfectants is yes | **386** |
| When Grocery is no & Vitamin is yes & Surface disinfectants is yes | **547** |
| When Grocery is yes & Taxi is no | **687** |
| When Grocery is yes & Vitamin is no & Taxi is yes & Face shield is no or not applicable | **741** |
| When Grocery is yes & Vitamin is yes & Taxi is yes & Children is no | **1013** |
| When Grocery is yes & Vitamin is no & Taxi is yes & Face shield is yes | **1073** |
| When Grocery is yes & Vitamin is yes & Taxi is yes & Children is yes | **1386** |

## Discussion

During COVID-19 period, a change in household spending has occurred to prevent the pandemic of COVID-19 for example: buying extra products such as masks, hand sanitizer, vitamins or changing the routine of life such as using taxi instead of public transportation or buying more food due to the long period of staying at home. Therefore, the aim of the current study is to assess the average extra costs of each items to predict the total average extra costs in household spending.

The present study reveals that, 11% of families had one person lost his job compared a survey conducted by Adams-Prassl in late March 2020 shows that 8% of workers having lost their job (7) and another study found that 3% of employee having lost their job (8). Regarding income, 62% of households reported reduction in total income (9) compared to 35.4% in the current study this difference may be due to that our respondents have high level of education (most of them are working in medical field) and those with lower levels of education are more likely to lose their jobs (9).

Regarding to the change of household spending, 49.8% of households had increase in spending (mean = 1215.6, SD = 560.8) while 14.8% had decrease in spending (mean = 1702.7, SD = 989.24). The reason for this decrease can clarify as that many activities had been stopped or delayed due to COVID-19 pandemic.

From Income, Expenditure and Consumption survey, the average annual expenditure of the family on food and drink in Egypt is 37.1%, 33.9% in urban, and 40.2% in rural of the total annual expenditure, which is the largest percentage of the family’s expenditure (10). The present study has similar results, the increasing in grocery spending was the dominated item during COVID-19 pandemic; has the maximum average cost change (mean = 707.2, SD = 530.7) compared to other items which can be understand that with more time spent at home, more food we eat which led to spend more money. Moreover, physicians stated that more people reported unexpected weight gain during the COVID-19 pandemic (11).

Concerning regression tree, the minimum average extra cost in household spending was 217 L.E while the maximum average extra cost was 1386 L.E, this result can be validated by comparing it with the change in household spending (min = 200 L.E, max= 2000 L.E).

## Limitation

Due to COVID-19 pandemic, face-to-face data collection was difficult to do, therefore snowball sampling technique was implemented to recruit individuals; highly educated only. Thus, this result can be generalized only among highly educated people.

## Recommendation

A phone survey is needed to be conducted to recruit all different groups in the community.

## Conclusion

The effect of COVID-19 pandemic in households spending varies largely between households, it depends on what they do to prevent COVID-19.

## Data Availability

All data are available from the corresponding author on responsible request.

## Ethical consideration

The study was approved by the Institutional Review Board and the Ethical Committee of High Institute of Public Health in Egypt. Anonymity and confidentiality of the participants’ data was ensured and maintained.

